# Accurate prediction of children’s target height from their mid-parental height

**DOI:** 10.1101/2022.10.31.22281712

**Authors:** Danny Zeevi, Adi Ben Yehuda, David Zangen, Leonid Kruglyak

## Abstract

**Background:** For the past 50 years, standard guidelines have recommended the use of sex-adjusted mid-parental height to predict a child’s final height. Here, we studied the accuracy of this procedure.

**Methods:** We used height data in a cohort of 23 very large nuclear families (Mean = 11 adult children per family). We compared the actual final height of the children to their height predicted by the standard procedure, as well as to alternative height predictions that incorporate corrections of mid-parental height for age, sex, and regression to the mean.

**Results:** Standard mid-parental height explained 36% of the variance in children’s heights, with a heritability of 74%, and children were on average 2.7 cm taller than predicted by their target heights. When we introduced a non-linear correction for the age of the parents, employed a multiplicative (rather than additive) correction for sex, and accounted for regression to the mean, the variance explained increased to 40%, heritability increased to 80%, and prediction bias was reduced from 2.7 cm to 0.14 cm (representing an improvement in prediction by half a standard deviation of the height distribution). We further measured the empirical distribution of heights of adult children around their predicted height. We describe how this distribution can be used to estimate the probability that a child’s height is within the normal expected range.

**Conclusions and Relevance:** Based on these observations, we propose an improved method for predicting children’s target heights. Our procedure for determining whether the deviation of a child’s projected height from the target height is in the normal range can be used to assess whether the child should be tested further for potential medical abnormalities.

## Introduction

Prediction of children’s target height from the heights of their parents is essential for estimating the children’s growth. Target height is defined as a child’s mid-parental height, adjusted for sex by adding 6.5 cm to predict a son’s height or subtracting 6.5 cm to predict a daughter’s height. Target height is typically compared to a child’s projected height, which is calculated by plotting the child’s current height on a standard growth chart and extrapolating to adult height at the same percentile. In the case of abnormal bone age, projected height is calculated using a bone atlas. A large discrepancy between projected and target heights is taken to indicate the possibility of an underlying medical condition. This procedure was developed by Tanner et al. nearly 50 years ago ^1^ and is still the recommended medical practice today ^2,3^.

However, the Tanner procedure has several problems that can lead to inaccurate and biased estimates of target height. When used in clinical practice, these inaccurate estimates may result in unintended errors in diagnostic decisions.

First, parental heights are usually not age-corrected. This leads to underestimation of target height because measured parental heights are typically shorter than past peak parental heights. Tanner did suggest a correction of adding 1.5 cm for parents between 45 to 55 years of age and 3 cm for parents above 55 years of age ^1^. However, longitudinal studies have since shown that height shrinkage starts around the age of 30 and that it is nonlinear with age ^4^. Furthermore, no correction for parental age is currently suggested in Pediatric Endocrinology guidelines ^2,3,5^.

Second, the Tanner correction of height for sex adds a constant 13 cm to transform female height to male height, regardless of the height itself. It is hard to envision a biological mechanism that would lead to a constant height difference between females and males, and it might be more accurate to use a correction proprotional to height or to standardize female and male heights using their means and standard deviations ^6^.

Third, mid-parental height is a biased predictor of children’s target height as a consequence of regression to the mean. Very tall parents tend to have shorter children than their mid-parental height predicts, while very short parents tend to have taller children. This observation was originally made over 130 years ago by Galton in his seminal work on heights in families ^7^. In fact, the term “regression” was first used in a statistical context in that paper.

Yet, regression is unaccounted for in the clinic today, despite previous suggestions to do so ^8^. Because of the directional nature of regression to the mean, ignoring it is expected to lead to systematic underdiagnosis of medically related short or tall stature.

Finally, the healthy normal range around the predicted target height is not well established. Tanner et al. used theoretical considerations to estimate that the child’s final adult height should fall within ±8.5 cm of mid-parental height, corresponding to the 3^rd^ and 97^th^ percentiles ^1^. They later adjusted this range to ±9 cm for girls and ±10 cm for boys ^9,10^. Others have suggested a 95% confidence interval of ±1.64 SDS ^10^, which under the CDC standards translates to ±10.6 cm for girls and ±11.7 cm for boys ^11^. The use of these cutoffs in the diagnosis of children with Idiopathic Short Stature (ISS) has been recommended by the Growth Hormone Research Society, the Lawson Wilkins Pediatric Endocrine Society, and the European Society for Paediatric Endocrinology ^5^. However, the use of rigid cutoffs to determine “normal” vs “abnormal” deviations from target height is arbitrary and may lead to misdiagnosis. The cutoffs are based on height measurements in young children who have not completed their growth. Furthermore, they come from studies of many small families and are not necessarily representative of the distribution of heights of adult children in a single nuclear family.

In order to address these limitations, we studied a unique cohort of 23 very large nuclear families with 7 to 16 (mean=11) adult children who have completed their growth. The large number of adult children per family allowed us to compare the target height predicted from mid-parental height to many measured final heights in every family. This enabled us to evaluate different target height estimators that incorporate the various corrections discussed above, as well as to measure the empirical distribution of heights of adult children around their predicted height.

Based on these observations, we propose an improved method for predicting children’s target heights. We also describe a procedure for determining whether the deviation of a child’s projected height from the target height is in the normal range or if the child should be tested further for medical abnormalities.

## Methods

### Recruitment and measurements of participants

Participants are a subset of a previous study of the genetic basis of height in large Jewish nuclear families ^12^. We included here all families for which we had heights of both parents. Participants were recruited in Israel and the US after obtaining IRB approval in both locations. Participants gave written informed consent, filled out a medical questionnaire, and had their height measured to the nearest 0.1 cm, with four technical replicates. The complete dataset is available at: https://www.ncbi.nlm.nih.gov/projects/gap/cgi-bin/study.cgi?study_id=phs001852.v1.p1

### Corrections of height for age, sex, and regression to the mean

We applied a nonlinear correction for age according to a previously published model ^12^. In short, the model captures shrinkage that starts at age 30 and accelerates at a constant rate, with different shrinkage coefficients for men and for women.

We tested three models for correction for sex:

1. Addition of a constant. We considered both 13 cm, as applied in the standard Tanner procedure, and 10.82 cm - the mean difference between males and females in our sample.
2. Multiplication of female heights by a constant. To determine the multiplication factor, we used heights from the CDC growth charts ^11^ and regressed male final height (age 20) on female height at the same percentile, while forcing the intercept to zero. This resulted in a multiplication factor (slope) of 1.08. Similar analysis on the heights in our cohort gave a multiplication factor of 1.066.
3. Standardization of male and female heights by Z-score. We tested this model with male and female means and standard deviations from both the CDC data and our sample.

We obtained equations for regression to the mean by regressing children’s measured heights on mid-parental heights. We tested all combinations of corrections for age and sex and calculated the resulting linear fit (**Table 1**).

**Table 1.** Performance of different schemes for correcting for age and sex. *Systematic error is calculated as the average difference between children’s final measured height and their target height.

| Correction for age | Correction for sex | Linear fit | Variance explained ($R^2$) | Heritability | Systematic error* |
| --- | --- | --- | --- | --- | --- |
| - | +13 cm | $Y=0.74X+47$ | 36% | 74% | 2.7 |
| + | +13 cm | $Y=0.79X+37$ | 37% | 79% | 0.06 |
| + | *1.08 | $Y=0.79X+36$ | 38% | 79% | 0.04 |
| + | Z score (CDC) | $Y=0.79X-0.077$ | 38% | 79% | 0.02 |
| + | +10.82 (Sample) | $Y=0.79X+36$ | 39% | 79% | 0.14 |
| + | *1.066 (Sample) | $Y=0.8X+35$ | 40% | 80% | 0.14 |
| + | Z score (Sample) | $Y=0.8X+0.017$ | 40% | 80% | 0.13 |

## Results

### Cohort details

We measured the final heights of 303 children and both parents from 23 large nuclear Jewish families with 7-16 adult children in each (mean=11). The age and height distributions are detailed in **Supplementary Table T1**. Average height of the children in our sample was within 1 cm of the average height of the Israeli population ^13^.

### Correction for age increases accuracy of target height prediction

To test the standard Tanner procedure, we corrected heights for sex by adding 13 cm to convert female height to male height, and regressed the height of children against their mid-parental height. Under this approach, mid-parental height explained 36% of the variance in the heights of children, and heritability of height was 74% (**Figure 1, Table 1**), similar to what has been observed in previous studies of other cohorts ^14^.

**Figure 1.**
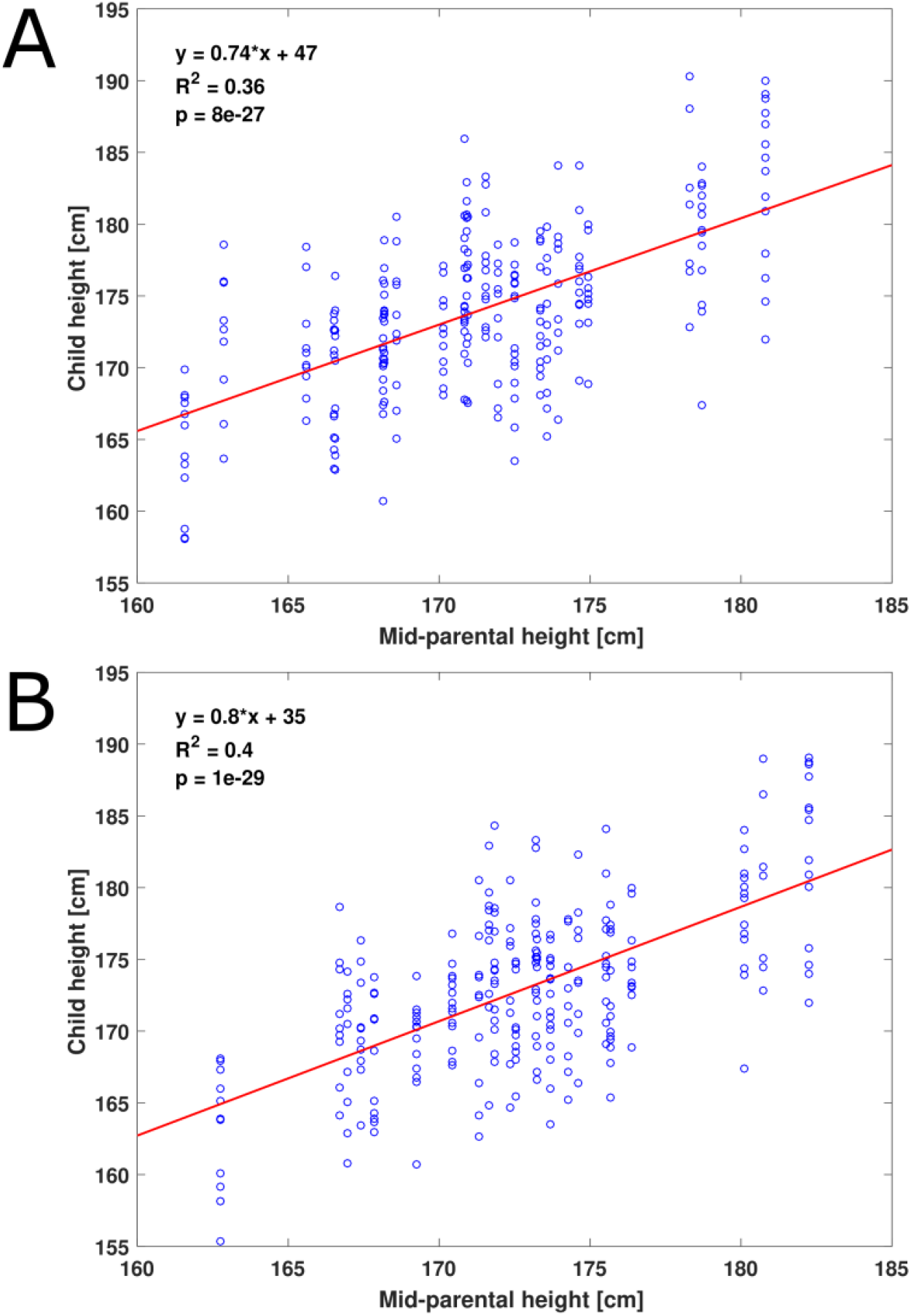
Variance in child height explained by mid-parental height. Each child’s height is plotted against the corresponding mid-parental height. Red line shows the linear fit. (**A**) Heights are corrected for sex by the standard Tanner procedure (adding 13 cm to female heights) and are not corrected for age. (**B**) Heights are first corrected for parental age and then corrected for sex by a multiplication factor.

To test the effect of parental age on the prediction of target height, we applied a nonlinear correction for age prior to the Tanner correction for sex. This step increased the variance explained to 37% and heritability to 79% (**Table 1)**.

More importantly, without the correction for age, the final heights of the children were on average 2.7 cm greater than their mid-parental height. After correction for age, this difference dropped to only 0.06 cm (**Table 1)**. This result suggests that in our cohort, most of the height difference between generations arises from the shrinkage of the parents rather than from a secular trend. Without correction for ages of the parents, children in the clinic may typically receive lower predicted target heights than the actual height they will reach, which may lead in turn to unnecessary medical intervention. It is important to note that our cohort shows an extreme example of this phenomenon because the parents in our cohort are 62±5 years old (mean±SD), older than typical parents in the clinic.

### Correction of height for sex is linearly dependent on height

The Tanner procedure is based on the assumption of a constant average 13 cm difference between female and male heights at the same percentile, independent of the actual percentile. To test whether this is the case, we investigated the height differences between adult males and females at the same percentile in the CDC growth charts (we used height at age 20, the final entry in the CDC growth charts) ^11^. We found that the difference between males and females is not constant, but rather is linearly dependent on height (R^2^=1, **Figure 2**).

**Figure 2.**
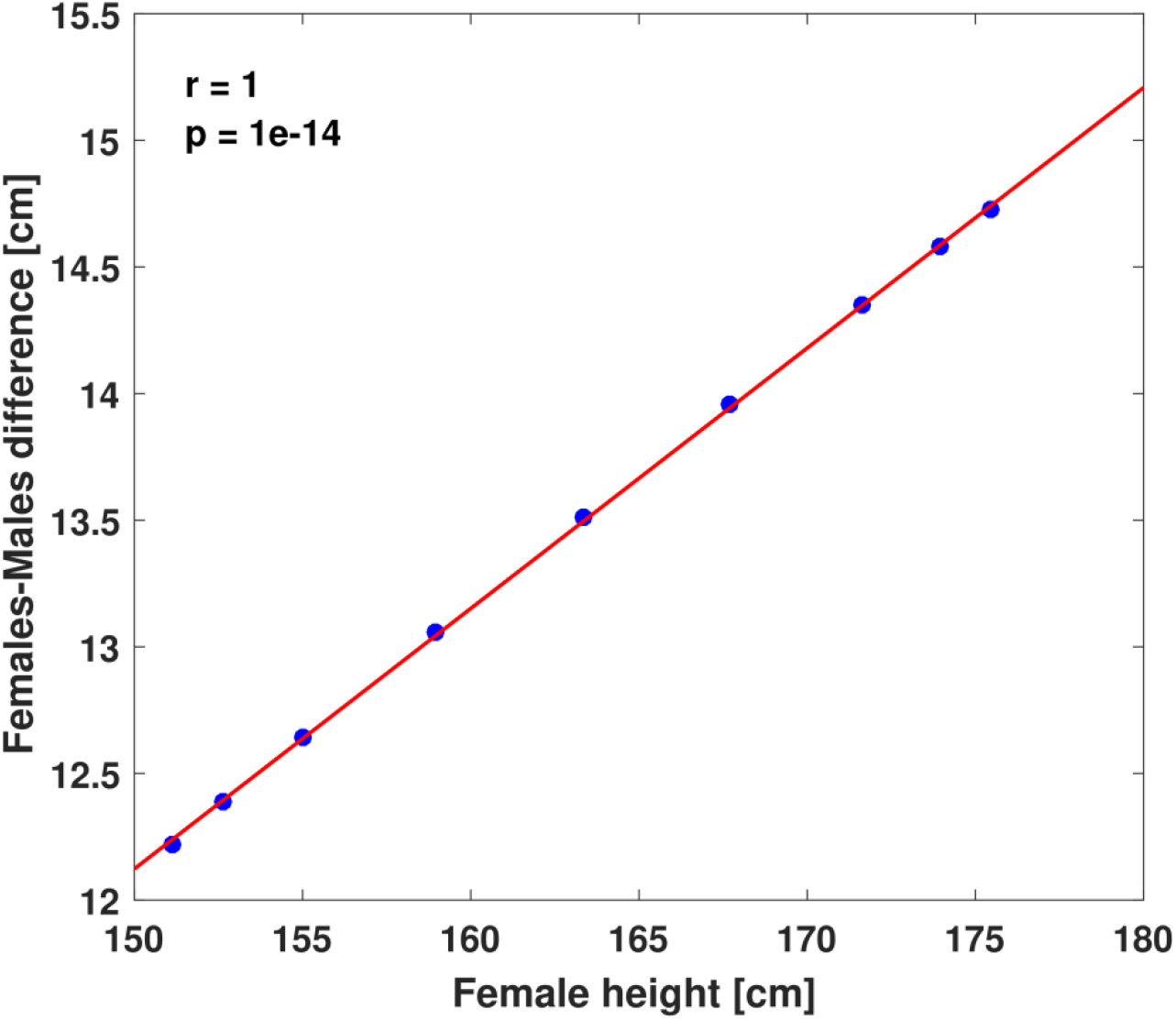
Height differences between males and females in the same percentile. Height differences between males and females in the same percentiles are plotted against female heights for the 3rd, 5th, 10th, 25th, 50th, 75th, 90th, 95th, and 97th percentiles. Heights are taken from the CDC growth charts ^11^. Red line shows the linear fit.

For example, while males in the 3^rd^ percentile are only 12.2 cm taller than females in the 3rd percentile, males in the 97^th^ percentile are 14.7 cm taller than females in the 97^th^ percentile. Fitting male heights to heights of females of the same percentile and forcing the intercept to zero showed a strong linear correlation, with Male height=1.08*Female height (R^2^=1, **Supplementary Figure S1**). Interestingly, multiplying female heights by 1.08 is the method that Galton used to correct height for sex in his 1886 publication ^7^. We corrected all heights for age and then for sex by multiplication by 1.08, and regressed the height of children against their mid-parental height. This resulted in a further increase of the variance explained by mid-parental height to 38% (**Table 1**). We tried a third method of standardizing the heights of females and males by Z-score. This resulted in R^2^=38%, similar to the multiplicative correction by 1.08.

Populations may vary in their patterns of male-female height differences. In our cohort, the mean difference between males and females was only 10.82 cm, and the multiplicative correction factor was 1.066. Cohort-specific constant correction by 10.82 cm increased the variance explained to 39%. Cohort-specific multiplicative correction by a factor of 1.066 further increased it to 40%, again favoring multiplicative over additive correction (**Figure 2**). We observed the same R^2^=40% for the Z-score standardization method that used the means and standard deviations of males and females in our cohort.

Although the overall improvement obtained by the use of a multiplicative correction for sex appears small (∼1% additional variance explained), it is important to note that the effect is most pronounced at height extremes, where medical relevance is greater than in the middle of the distribution. For example, correcting a father’s height in the 97^th^ percentile by subtracting 13 cm will result in an error of 1.7 cm in a daughter’s predicted height, while correcting it by dividing by 1.08 reduces the error to only 0.6 cm. In contrast, for a father’s height in the 50^th^ percentile, the corresponding errors will be 0.5 cm and 0.4 cm.

### Accounting for regression to the mean increases the accuracy of height prediction

To assess the effect of regression to the mean on height prediction, we calculated the difference between the mean height of children in each nuclear family and their mid-parental height. As expected, we observed a negative correlation (r=-0.47, P=0.02, **Figure 3**). For standardized heights, we observe the following relationship: Corrected Height = 0.79*(mid-parental standardized height) – 0.077 (**Table 1**).

**Figure 3.**
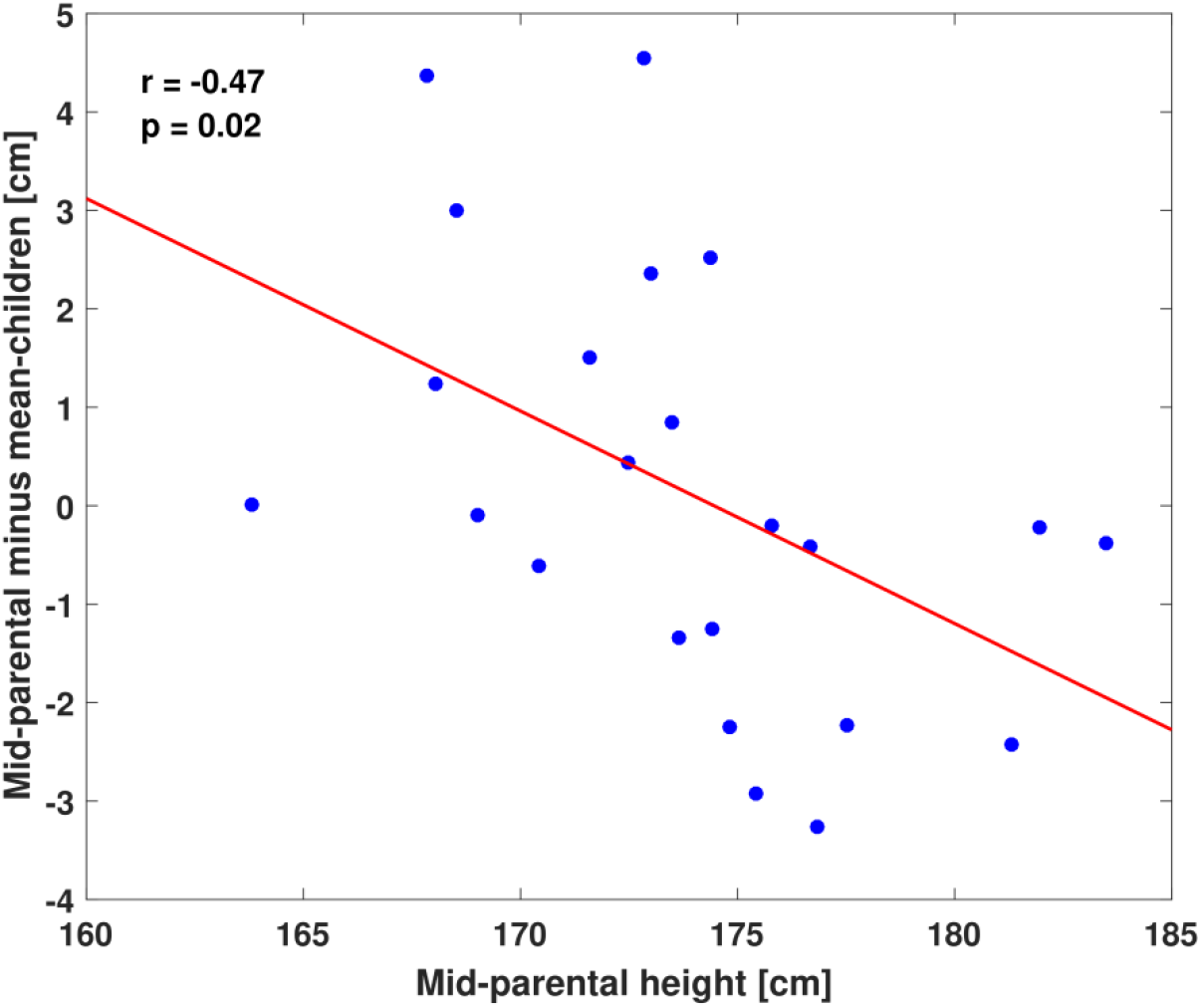
Regression to the mean. For each family, the difference between mid-parental height and mean height of the children is plotted against the the mid-parental height. Red line shows the linear fit. Tall parents tend to have shorter children than their mid-parental height would predict, while short parents tend to have taller children. The correction for sex used for this plot is multiplication by 1.08, but results are similar for other correction schemes.

The effect of regression to the mean is larger and more relevant for very tall or very short parents, who are also more likely to come to the clinic. For example, short parents who are both in the 3^rd^ percentile would be predicted to have children in the 3^rd^ percentile if the standard guidelines are applied, but in the 6^th^ percentile, 2-3 cm taller than their parents, if regression to the mean is accounted for as we suggest. Therefore, not accounting for regression to the mean will result in underestimating the final height of short children and overestimating the final height of tall children, potentially leading to errors in medical decisions on both ends of the height spectrum.

### Distribution of children’s height around their target height

The large sizes of the families in our cohort allowed us to investigate, at the single family level, the distribution of children’s height around their target height (mid-parental height corrected for age, sex and regression to the mean). We find that the standard deviation of the residuals around the target height within a family is on average 4.5±0.9 cm for sons and 4.2±0.8 cm for daughters (Mean over all families±SD, with sex correction via multiplication by 1.08). We do not observe a significantly wider distribution of the residuals for taller parents (r=0.38, p=0.08, **Supplementary Figure S2**), similar to previous findings in a population-based study ^15^.

When all the children from all families are combined, the standard deviation of the residuals is slightly higher: 4.7 cm for sons and 4.4 cm for daughters. Taken together, these results suggest that the distributions of children’s heights around their mid-parental height are similar between different families, and that the means and standard deviations from our sample can be used in the clinic for other families.

Under the assumption of a normal distribution, we can calculate the probability that a child’s height will differ by any given amount from the mid-parental target height (**Supplementary Methods**). For example, the probability that a child will be at least 10 cm shorter than the target height is 1.5% for boys and 1% for girls. For a child who comes to the clinic, such a calculation can be used as an approximate estimate of the probability that the current height is not due to a medical condition, but simply falls within the expected distribution around the target height. This approach may be preferable to the arbitrary cutoffs that are currently in use.

## Discussion

For the past 50 years, the Tanner procedure has been the standard of care for determining if a child’s current height deviates from the expected height, necessitating further testing for medical abnormalities. Here, by studying large families with healthy children who completed their growth, we quantify inaccuracies in the Tanner procedure that could lead to suboptimal medical decisions. We show that not correcting parental height for age, correcting height for sex by adding a constant, and ignoring regression to the mean all lead to errors in estimating the target height of a child from the current heights of the parents. We further suggest the use of our empirical distribution of heights of adult children to calculate the probability that a child is indeed within the expected normal range of height.

For example, consider a case in which two parents who are one standard deviation below the mean (170 cm and 157 cm for the mother and the father, respectively) come to the clinic with a 9 years old girl who is short - 122 cm, in the 3^rd^ percentile with bone age similar to chronological age. Mid-parental height using the Tanner procedure is 157 cm (after the father’s height is reduced by 13 cm to correct for sex), and the predcited height of the daughter (3^rd^ percentile at age 20) is 151 cm, only 6 cm shorter than her mid-parental height. However, if one corrects for the ages of the parents (e.g., 45 and 50), corrects for sex by dividing the father’s height by 1.08 or by standardizing heights by Z-score according to the CDC parameters, and accounts for regression to the mean, the predicted height is actually 159 cm, and the difference between the target and projected heights is 8 cm. While this difference is within the “normal” limits defined by Tanner (±9 cm), comparing this difference to the normal distribution that we observe in our large families suggests that the probability of a difference this large occuring by chance is only 3%, indicating a greater necessity for tests and further evaluation.

A similar situation arises if the father is short (163 cm, 3rd percentile for men) while the mother’s height is average (163 cm, 50^th^ percentile for women). While it is easy to assume in this case that the girl’s short stature is inherited from her father, the actual probability of the daughter being that short is, according to our empirical distribution, only 3%, even when the father’s height is taken into account.

To facilitate the application of our suggested method, we supply in the **Supplementary Methods** a detailed algorithm to correct parental heights for age and sex, compute the child’s target height accounting for regression to the mean, and calculate the probability that the deviation between the target and projected height is within the expected normal range. This algorithm is ready for implementation in software packages for height prediction.

In the future, this algorithm could be improved further by inclusion of genotype information ^16^ ; we will explore this possibility in a future study.

## Data Availability

All data produced in the present study are available online at: https://www.ncbi.nlm.nih.gov/projects/gap/cgi-bin/study.cgi?study_id=phs001852.v1.p1

## Author contribution

All authors made substantial contributions to conception and design, acquisition of data, or analysis and interpretation of data.

Conceptualization - D.Ze., D.Za. and L.K.; Methodology - D.Ze. and D.Za.; Formal Analysis - D.Ze.; Investigation - D.Ze.; IRB application - D.Ze. and A.B.; Data Curation - D.Ze.; Writing, original draft preparation - D.Ze.; Writing, review & editing - D.Ze., A.B., D.Za. and L.K.; Visualization - D.Ze.; Supervision - L.K.; Project Administration - A.B.; Funding Acquisition - L.K.

