## Supplementary Material for "Accurate prediction of children’s target height from their mid-parental height"

### Supplementary Methods

**Supplementary Table T1.** Mean height and age of participants

**Supplementary Figure S1.** Mean heights of males and females in the same percentiles

**Supplementary Figure S2.** Height differences between target and actual heights of children in different families

### Supplementary References

**Supplementary Methods**

**Complete procedure for calculating the target and projected heights of a child, and for deducing the probability that a deviation between the two is abnormal**

Use centimeters for all following calculations:

1) Measure accurately the height of the child and his parents.

2) Correct height of parents for age according to the previously published model [1]:

$$X \equiv Height\begin{pmatrix} age \\ corrected \end{pmatrix} = \begin{cases} Height, & Age \leq 30 \\ Height - 0.00208 \times (Age-30)^2, & Males, Age > 30 \\ Height - 0.003205 \times (Age-30)^2, & Females, Age > 30 \end{cases}$$

3) Correct height for sex by standardizing (z-scoring) parents and child's heights according to the L,M,S parameters in the CDC growth chart [2], available at https://www.cdc.gov/growthcharts/data/zscore/statage.xls

Assuming L≠0:

$$Z = \frac{\left(\frac{X}{M}\right)^L - 1}{L * S}$$

Where:

$Z$ ≡ Standardized height. Calculate $Z_{child}, Z_{father}, Z_{mother}$, each according to the relevant X, M and S parameters in the CDC data for specific sex and age.

$X$ = Measured height in cm (after correction for age if it is a parent)

$M$ = Median for age according to CDC data for the relevant age and sex. Use $M$ at age 20 (last entry) for parents.

$S$ = Generalized coefficient of variation according to CDC data for the relevant age and sex. Use $S$ at age 20 (last entry) for parents.

$L =$ Power in the Box-Cox transformation according to CDC data for the relevant age and sex. Use $L$ at age 20 (last entry) for parents.

If working with a specific population that the CDC data does not fit well, and this population's means and standard deviations are known for all ages, then standardize by:

$$Z = \frac{X - \overline{X}}{\sigma}$$

Where:

$X =$ Measured height in cm (after correction for age if it's a parent)

$\overline{X} =$ Mean height for the specific population and age

$\sigma =$ Standard deviation height for the specific population and age

4) Define

$$Z_{projected} \equiv Z_{child}$$

Where:

$Z_{projected} \equiv$ Projected (standardized) height of the child in adulthood given by his current standardized height (assuming that the child will be at the same percentile when he completes his growth)

5) Calculate mid-parental height

$$Z_{midparental} = \frac{Z_{father} + Z_{mother}}{2}$$

6) Correct mid-parental height for regression to the mean, to increase the accuracy of the prediction of height of the child:

$$Z_{target} = 0.79 * Z_{midparental} - 0.077$$

Where:

$Z_{target} \equiv$ Target (standardized) height of the child in adulthood according to his parents' heights.

The parameters for the slope and intercept were taken from the linear fit of plotting $Z_{child}$ against $Z_{midparental}$

7) In order to get results in cm, transfer standardized projected heights and standardized target heights to cm according to the CDC parameters:

$$H_{projected} = M_{20} * (1 + L_{20} * S_{20} * Z_{projected})^{\frac{1}{L_{20}}}$$

$$H_{target} = M_{20} * (1 + L_{20} * S_{20} * Z_{target})^{\frac{1}{L_{20}}}$$

Where:

$H_{proj} \equiv$ Projected height of the child in cm

$H_{target} \equiv$ Target height of the child in cm

$M_{20}$ = CDC median height at age 20 and for sex same as the child's

$S_{20}$ = CDC Generalized coefficient of variation at age 20 and for sex same as the child's

$L_{20}$ = CDC Power in the Box-Cox transformation at age 20 and for sex same as the child's

If working with a specific population that the CDC data does not fit well, and this population's means and standard deviations are known for all ages, then transform by:

$$H_{projected} = Z_{projected} * \sigma_{adult} + \overline{X}_{adult}$$

$$H_{target} = Z_{target} * \sigma_{adult} + \overline{X}_{adult}$$

Where:

$\overline{X}_{adult}$ = Mean height of adults in the specific population, for sex same as the child's

$\sigma_{adult}$ = Standard deviation of height of adults in the specific population for sex same as the child's

8) Compare the child's projected height to his target height and calculate the probability that the difference occurred by chance:

$$\Delta H = H_{projected} - H_{target}$$

For short children, the probability that their projected height is by chance shorter than their target height by $\Delta H$ or more:

$$P\left(H \leq H_{projected} \middle| H_{target}\right) = \frac{1}{\sigma_{fam}\sqrt{2\pi}} \int_{-\infty}^{H_{projected}} e^{\frac{-(t-H_{target})^2}{2(\sigma_{fam})^2}} dt$$

For the calculation, one can simply use the Matlab function:

$$P\left(H \leq H_{projected} \middle| H_{target}\right) = normcdf(H_{projected}, H_{target}, \sigma_{fam})$$

For tall children:

$$P\left(H \geq H_{projected} \middle| H_{target}\right) = \frac{1}{\sigma_{fam}\sqrt{2\pi}} \int_{H_{projected}}^{\infty} e^{\frac{-(H_{target}-t)^2}{2(\sigma_{fam})^2}} dt$$

In Matlab:

$$P\left(H \geq H_{projected} \middle| H_{target}\right) = normcdf(H_{projected}, H_{target}, \sigma_{fam}, 'upper')$$

Where:

$\sigma_{fam}$= Standard deviation of height of adult siblings within a family, according to the sex of the child. Our large families data suggest that:

$\sigma_{fam}(male\ siblings) = 4.6$ cm

$\sigma_{fam}(female\ siblings) = 4.2$ cm

**Supplementary Table T1. Mean height and age of participants**

| Participants | Number of participants | Age [yrs] (Mean±SD) | Height [cm] (Mean±SD) |
|---|---|---|---|
| Fathers | 23 | 64±5 | 169.9±5.9 |
| Mothers | 23 | 60±4 | 159.3±5.9 |
| Sons | 139 | 30±6 | 172.9±5.9 |
| Daughters | 118 | 30±7 | 162±5.5 |

**Supplementary Figure S1. Mean heights of males and females in the same percentiles**

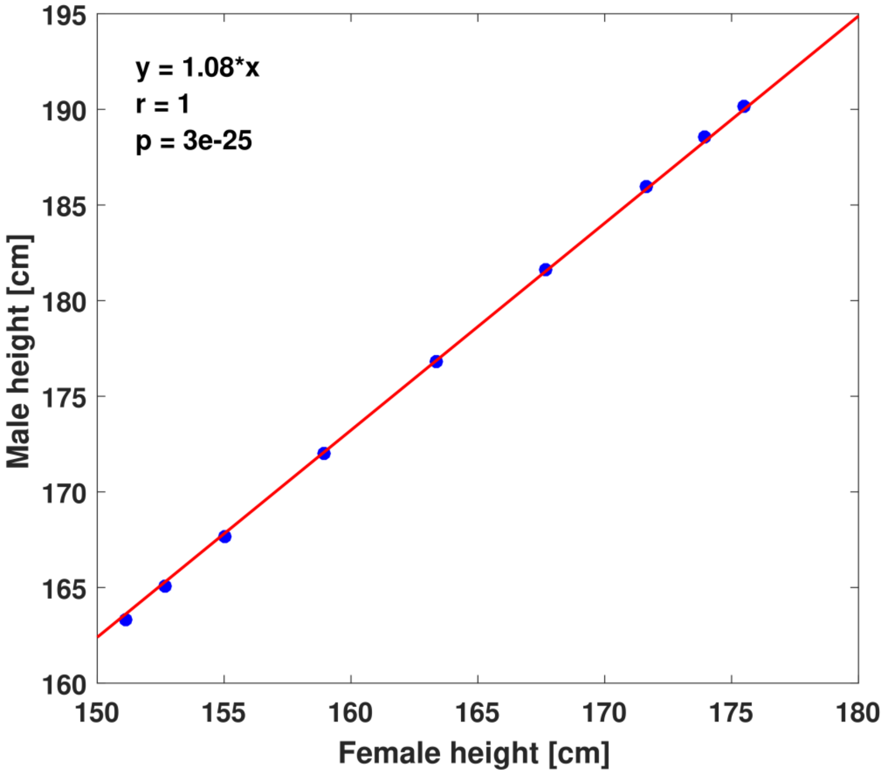


Male mean heights of the 3rd, 5th, 10th, 25th, 50th, 75th, 90th, 95th, and 97th percentiles plotted against female mean heights of the same percentiles. Height data is from the CDC growth charts for age 20 [2]. Red line shows the linear fit.

**Supplementary Figure S2. Height differences between target and actual heights of children in different families**

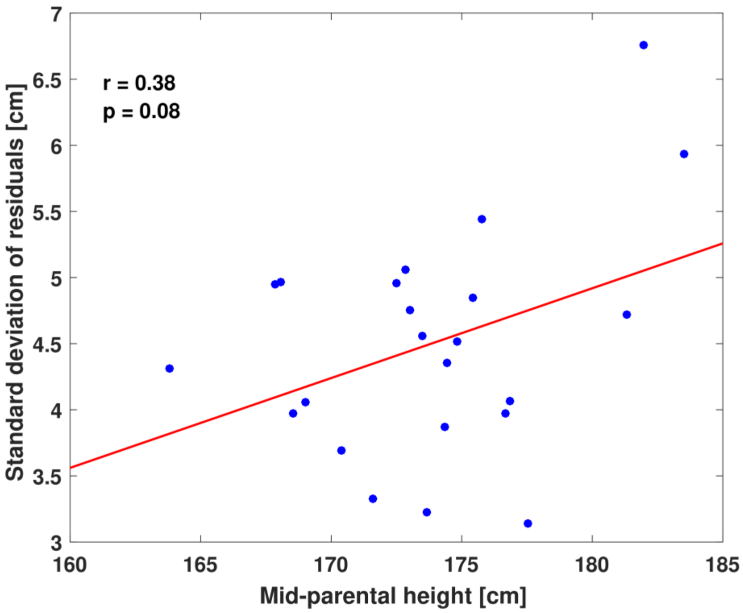


For each family, the standard deviation of the differences between the actual heights of the children in that family and their target height as calculated from their parents (mid-parental height corrected for age, sex and regression to the mean). Red line shows the linear fit.
